# Summer and SERT: Effect of daily sunshine hours on *SLC6A4* promoter methylation in seasonal affective disorder

**DOI:** 10.1101/2024.10.25.24316134

**Authors:** P. A. Handschuh, M. Murgaš, D. Winkler, E. Winkler-Pjrek, A. M. Hartmann, K. Domschke, P. Baldinger-Melich, D. Rujescu, R. Lanzenberger, M. Spies

## Abstract

Meteorological factors affect the serotonergic system, potentially influencing *SLC6A4* promoter methylation in seasonal affective disorder (SAD). However, studies on how sunlight impacts methylation and modulates SERT activity in this context remain limited. This study aims to investigate the effect of average daily sunshine duration on *SLC6A4* promoter methylation in a cohort consisting of both patients with SAD as well as healthy controls (HC). Methylation data were collected from 28 patients with SAD and 40 healthy controls (HC). Average methylation from four *SLC6A4* promoter CpG sites was assessed. Daily sunlight data for Vienna, Austria (mean of 28 days before blood sampling), were obtained from ©GeoSphere Austria. A general linear model (GLM) analyzed *SLC6A4* promoter methylation as the dependent variable, with sunlight hours as the independent variable, and group (SAD, HC), age, sex, and 5-HTTLPR/rs25531 as covariates. Exploratory analyses examined sunlight hours and methylation effects on Beck Depression Inventory (BDI) scores. Sunlight had a significant effect on *SLC6A4* promoter methylation (p = 0.03), with more sunlight hours resulting in lower methylation (r = −0.25). However, the interaction between sunlight and group was non-significant, suggesting a rather general effect across both groups. Sunlight also influenced BDI scores (p < 0.01), with fewer sunlight hours leading to higher BDI scores (r = −0.25), which aligns with previous research. *SLC6A4* promoter methylation had no significant effect on BDI scores. Our findings suggest that sunlight impacts *SLC6A4* promoter methylation, but this effect appears general, not specific to SAD pathophysiology.

## INTRODUCTION

The relevance of the serotonin transporter (SERT) to the pathophysiology and treatment of various psychiatric disorders is well established. Changes to SERT density were shown in depression and anxiety in humans *in vivo*. The SERT is a central target of various psychopharmacologic treatments, first and foremost selective serotonin reuptake inhibitors (Spies, Knudsen et al. 2015).

The serotonin transporter gene (*SLC6A4*) length polymorphism (5-HTTLPR) and the associated rs25531 variant are likely among the most studied serotonergic gene variants in the context of psychiatric disorders (Caspi, Sugden et al. 2003). Epigenetic regulation, such as DNA methylation of the *SLC6A4* gene, was shown to mediate risk for psychiatric disorders. For example, peripheral *SLC6A4* promoter methylation was linked to depression (Kim, Stewart et al. 2013, Booij, Szyf et al. 2015, Iga, Watanabe et al. 2016, Schiele, Kollert et al. 2019) as well as related structural (Dannlowski, Kugel et al. 2014, Booij, Szyf et al. 2015, Won, Choi et al. 2016) and functional (Frodl, Szyf et al. 2015, Ismaylova, Levesque et al. 2018, Schneider, Kugel et al. 2018) imaging endophenotypes. In addition, methylation status may be related to treatment outcomes (Domschke, Tidow et al. 2014, Okada, Morinobu et al. 2014, Schiele, Zwanzger et al. 2021). Various stress-related risk factors for affective disorders were shown to result in altered *SLC6A4* promoter methylation, with both increases (Zhao, Goldberg et al. 2013, Booij, Szyf et al. 2015, Duman and Canli 2015) and decreases reported (Devlin, Brain et al. 2010, Alasaari, Lagus et al. 2012). Thus, changes to *SLC6A4* promoter methylation may serve as an intermediate between disease risk and the clinical manifestation of psychiatric illnesses.

The serotonergic system is susceptible to season (Matheson, Schain et al. 2015) and light (Spindelegger, Stein et al. 2012, Harrison, Tyrer et al. 2015, Tyrer, Levitan et al. 2016). Cerebral SERT protein levels are higher in fall/winter and lower in spring/summer (Ruhe, Booij et al. 2009), and show a negative association with sunshine levels (Praschak-Rieder, Willeit et al. 2008). Dysregulation of seasonal fluctuations in SERT expression may underly seasonal affective disorder (SAD) (Mc Mahon, Andersen et al. 2016), which is characterized by depressive symptoms in fall/winter and remission in spring/summer, and for which season and light are both risk factors and directly impact on pathophysiology (Mersch, Middendorp et al. 1999). In fact, a reduction in brain SERT levels was shown after bright light therapy (BLT), the gold-standard treatment for SAD (Harrison, Tyrer et al. 2015, Tyrer, Levitan et al. 2016), underlining the SERT’s pathophysiologic relevance.

Various environmental factors impact epigenetic processes within the brain (Lim 2021). Despite the potentially pronounced pathophysiologic relevance of this relationship, little is known about the effect of season on epigenetic regulation of the serotonin system in general, and the SERT in particular. In theory, alterations in methylation mediated by seasonal and environmental factors might underlie the changes in SERT expression observed in SAD. Of seasonal variables, temperature (Bind, Zanobetti et al. 2014, Xu, Li et al. 2021) and light exposure (Da Silva Melo, Barroso et al. 2015, Vandiver, Irizarry et al. 2015) were shown to influence methylation patterns in the human genome. However, the influence of these environmental factors on *SLC6A4* promoter methylation has yet to be elucidated.

Here, we aimed to investigate this relationship by assessing the impact of meteorologic measures, gleaned from regional assessments, on peripheral *SLC6A4* promoter methylation in a cohort of patients with SAD as well as healthy controls (HC). We analyzed methylation within four *SLC6A4* promoter CpG sites previously linked to affective disorders and their treatment (Domschke, Tidow et al. 2014).

## EXPERIMENTAL PROCEDURES

### Study Design

For the study at hand, methylation data from 28 (18/10 f/m) patients diagnosed with SAD and 40 HC (23/17 f/m) were analyzed (n = 68). Data were gleaned from a previously published placebo-controlled study using positron emission tomography (PET) to investigate the effect of BLT on MAO-A density in the brain across the seasons (Spies, James et al. 2018). This previous study encompassed a screening visit, a structural magnetic resonance imaging (MRI) scan, 3 PET scans (PET1, before BLT-treatment in autumn or winter; PET2, after BLT-treatment in autumn/winter; and PET3, after BLT-treatment in spring or summer) as well as a follow-up visit. For the analyses at hand, no imaging data (i.e., PET or MRI data) was used. Blood draw for methylation analysis was either conducted before PET1 or PET2 in autumn or winter (September to February) or before PET3 in spring or summer (March to August). Beck Depression Inventory (BDI) (Beck, Ward et al. 1961) scores were assessed in the same season as the blood sample was taken. If the BDI score was obtained in a different season, the patient was excluded from BDI-related analyses. The study was conducted in accordance with the Declaration of Helsinki, taking all current revisions and the good scientific practice guidelines of the Medical University of Vienna into account. The protocol was approved by the ethics committee of the Medical University of Vienna (EK Nr.: 1681/2016) and registered at clinicaltrials.gov (NCT02582398).

### Participants

Patients with SAD were enrolled at the outpatient clinic of the Department of Psychiatry and Psychotherapy at the Medical University of Vienna. HC were recruited via dedicated message boards at the Medical University of Vienna. The Structured Clinical Interview for DSM-4 Axis I disorders (SCID-I) (First 2002) as well as the Seasonal Pattern Assessment Questionnaire (SPAQ) (Rosenthal 1984) were performed in all patients to confirm the diagnosis of unipolar winter-type SAD and to exclude any other type of psychiatric condition. The SCID-I and the SPAQ were used in HC to exclude psychiatric diagnoses, particularly SAD. Participants were excluded if they were undergoing current psychopharmacological treatment or had received such treatment within six months prior to enrolment. Additional exclusion criteria included severe somatic or neurologic comorbidities, current smoking, drug abuse, pregnancy, or lactation. A trained physician assessed potential contraindications during the initial visit through a review of medical history, routine laboratory tests (blood and urine), electrocardiography, and a physical examination. All participants provided written informed consent and received financial compensation for their participation.

### Methylation Analysis and *SLC6A4* Genotyping

DNA methylation analysis was performed at the Department of Psychiatry and Psychotherapy at the University of Freiburg, Faculty of Medicine, Germany via direct sequencing of bisulfite-converted DNA. Methylation levels at 4 CpG sites (CpG1 = 30,236,072; CpG2 = 30,236,084; CpG3 = 30,236,089; CpG4 = 30,236,091) (Domschke, Tidow et al. 2014) located in a 20 bp amplicon in the *SLC6A4* promoter (chromosome 17: 30,236,072–30,236,091; GRCh38.p2 Primary Assembly, UCSC Genome Browser) were analyzed in every individual sample by pyrosequencing (PyroMark Q96 ID, Qiagen) as described previously (Ziegler, Richter et al. 2016). All subjects were genotyped for the *SLC6A4* promoter repeat length polymorphism (5-HTTLPR), as the short allele of 5-HTTLPR has been associated with depression (Collier, Stöber et al. 1996), SAD (Rosenthal, Mazzanti et al. 1998) and seasonality (Johansson, Willeit et al. 2003). Additionally, genotyping included the *SLC6A4* single nucleotide polymorphism (SNP) rs25531, which modifies the long allele (L) of 5-HTTLPR. The rs25531 polymorphism differentiates between the LA variant (which retains high serotonin transporter expression) and the LG variant (which reduces serotonin transporter expression and is functionally similar to the short allele, S) (Kraft, Slager et al. 2005, Hu, Lipsky et al. 2006). Thus, in this study, S1 refers to both the short allele (S) and the LG variant of the long allele, as both are functionally similar in their effects on serotonin transporter expression. Genotypes were categorized into three groups: S1+S1, S1+LA, and LA+LA. Genotyping was performed at the Department of Psychiatry, Psychotherapy and Psychosomatics of the University of Halle, Germany, as outlined in Baldinger et al. (Baldinger, Kraus et al. 2015).

### Assessment of Meteorological Data

Datasets on daily sunlight hours and temperature for the 28 days preceding blood sampling were obtained for each participant from two separate meteorological measurement stations in Vienna, Austria. These data were sourced from © GeoSphere Austria (https://data.hub.geosphere.at). The first station is located in the outskirts of Vienna (“Hohe Warte”), while the second is situated in the city center (“Innere Stadt”). For each individual, the meteorological data from both stations were averaged. These averaged values were then utilized for subsequent statistical analyses. Due to the high correlation (r=0.86) between average sunlight hours and average temperature, only sunlight hours were considered in the final statistical procedures.

### Statistical Analysis

Statistical tests were performed using SPSS version 28 for Windows (SPSS Inc., Chicago, IL, USA).

#### Analyses of SLC6A4 Promoter Methylation

Average *SLC6A4* promoter methylation was described as the average methylation of all pre-defined CpG sites (CpG 1-4) and could be obtained from 40 (23/17 females/males) HC (mean age 34.35 ± SD 9.96 years) and 28 (18/10 females/males) patients with SAD (mean age 32.86 ± SD 9.55 years). In a first step, average CpG methylation was tested for normality using the Shapiro-Wilk test (p = 0.54; normally distributed). A general linear model (GLM) was then applied to investigate the relationship between average CpG methylation (dependent variable) and average sunlight hours (independent variable). The model also included the covariates group (patients with SAD vs. HC), 5-HTTLPR/rs25531 genotype, sex, and age. All continuous variables (i.e., average CpG methylation, age, average sunlight hours) were z-scored. Then, a second GLM was calculated in an exploratory manner with the interaction effect between group and average sunlight hours as an additional covariate to investigate whether there is a pathophysiologically relevant effect of SAD. Post hoc analyses were performed using Spearman’s correlation.

#### Beck Depression Inventory

BDI scores were collected and incorporated into the study to quantify the extent of depressive symptoms and to explore the potential relationship between this clinical parameter, sunlight exposure, and *SLC6A4* promoter methylation.

BDI scores were available for 22 (13/9 females/males) patients with SAD (mean age 31.41 ± SD 9.67 years) and 28 (16/12 females/males) HC (mean age 33.18 ± SD 9.9 years). To investigate the relationship between the BDI scores (dependent variable) and average sunlight hours (independent variable) in patients with SAD, a GLM with the covariates 5-HTTLPR/rs25531 genotype, sex and age was calculated. The same model was applied using the BDI scores of HC. Of note, the analysis was split due to the naturally high difference in BDI between patients with SAD and HC.

In a second BDI-related GLM, the potential influence of *SLC6A4* promoter methylation (independent variable) on BDI scores (dependent variable) was calculated in patients with SAD, using 5-HTTLPR/rs25531 genotype, sex and age as covariates. Again, the model was repeated using the BDI scores of HC. In both models, all continuous variables (i.e., BDI scores, average CpG methylation, age, average sunlight hours) were z-scored. The reduced sample size for this model can be attributed to the criterium limiting participants to those whose BDI scores were assessed in temporal proximity (e.g., in the same season) with the blood draws for methylation analysis. A correction for the number of subject groups (SAD patients and HC) was performed via Bonferroni correction, significance was set at p < 0.05.

## RESULTS

### Analyses of *SLC6A4* Promoter Methylation

As shown in *Figure 1*, the main model revealed a significant influence of average sunlight hours on average promoter methylation (p = 0.03). This could be confirmed by post hoc Spearman’s correlation analysis (r = −0.25), indicating that more sunlight hours result in lower methylation levels. Regarding the covariates included in the model, no significant effect of group (patients with SAD vs. HC), 5-HTTLPR/rs25531 genotype, sex, or age was found.

**Figure 1:**
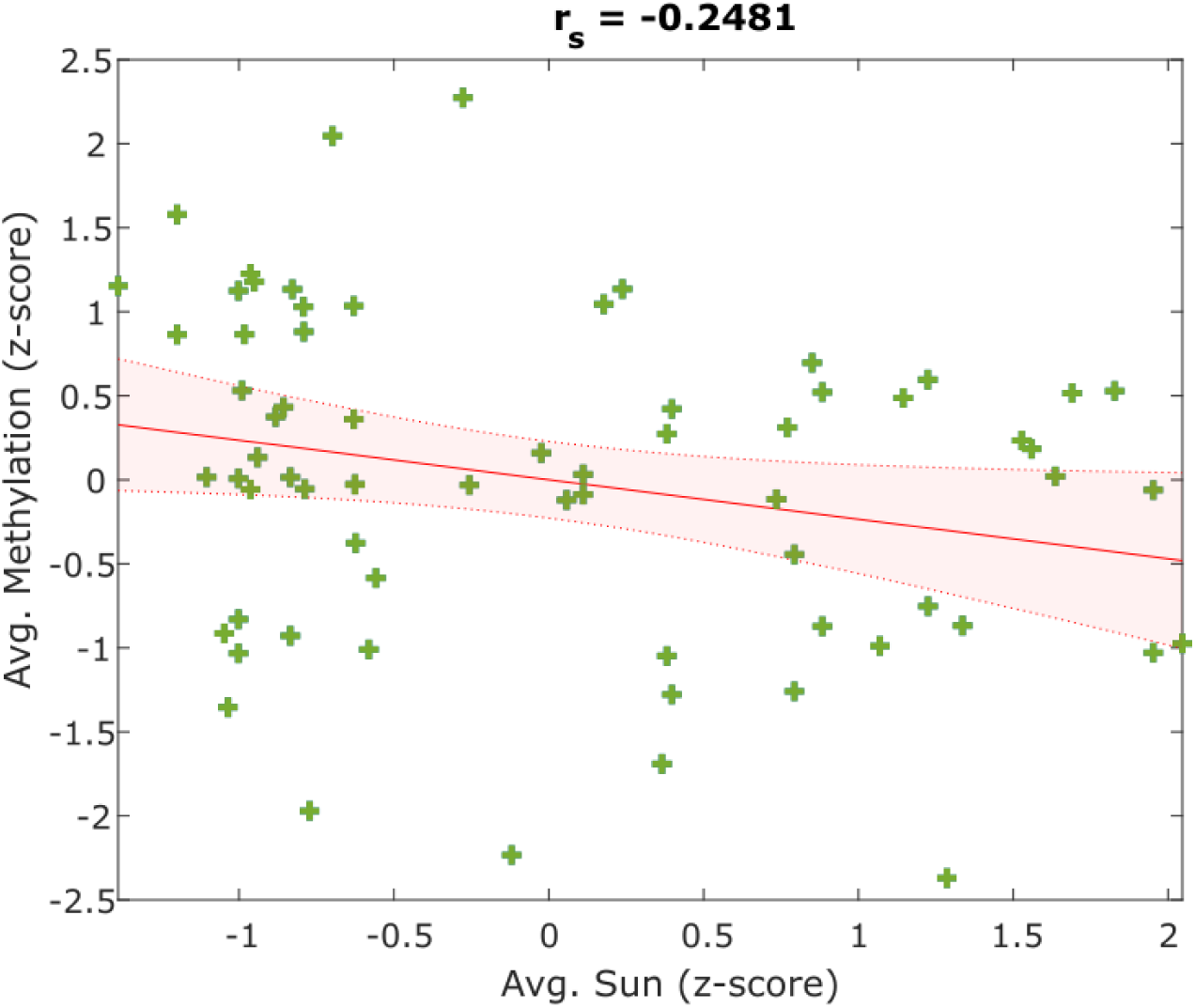
The plot illustrates the significant influence of sunlight hours per day, averaged from a time frame of 28 days prior to blood sampling, on average *SLC6A4* promoter methylation (p_uncorr_. = 0.03). Values are z-scored. The main model shows a negative correlation, confirmed by post hoc Spearman’s correlation analysis (r = −0.25), indicating that increased sunlight hours are associated with lower methylation levels of the *SLC6A4* promoter.

Adding the interaction between average sunlight and group as a potential covariate to the model (exploratory GLM model) resulted in a non-significant outcome between this factor and average sunlight hours (p = 0.7).

### Beck Depression Inventory

The exploratory analysis investigating the potential impact of average sunlight hours on BDI scores in patients with SAD yielded a significant finding; as demonstrated in *Figure 2*, an increase in average sunlight hours resulted in a reduction in BDI scores (p < 0.01). This could be confirmed by post hoc Spearman’s correlation analysis (r = - 0.41). No significant relationship between average sunlight hours and BDI scores was observed in HC. Although the model indicated a significant effect of gender on BDI scores, with women (p_uncorr._ = 0.04) showing higher BDI scores than men within the subpopulation of patients with SAD, this finding did not remain significant after correction for multiple comparisons. Therefore, this result should be interpreted with caution and may not represent a true effect. Additionally, the model investigating the influence of average *SLC6A4* promoter methylation on BDI scores yielded no significant results in neither the SAD nor the HC group.

**Figure 2:**
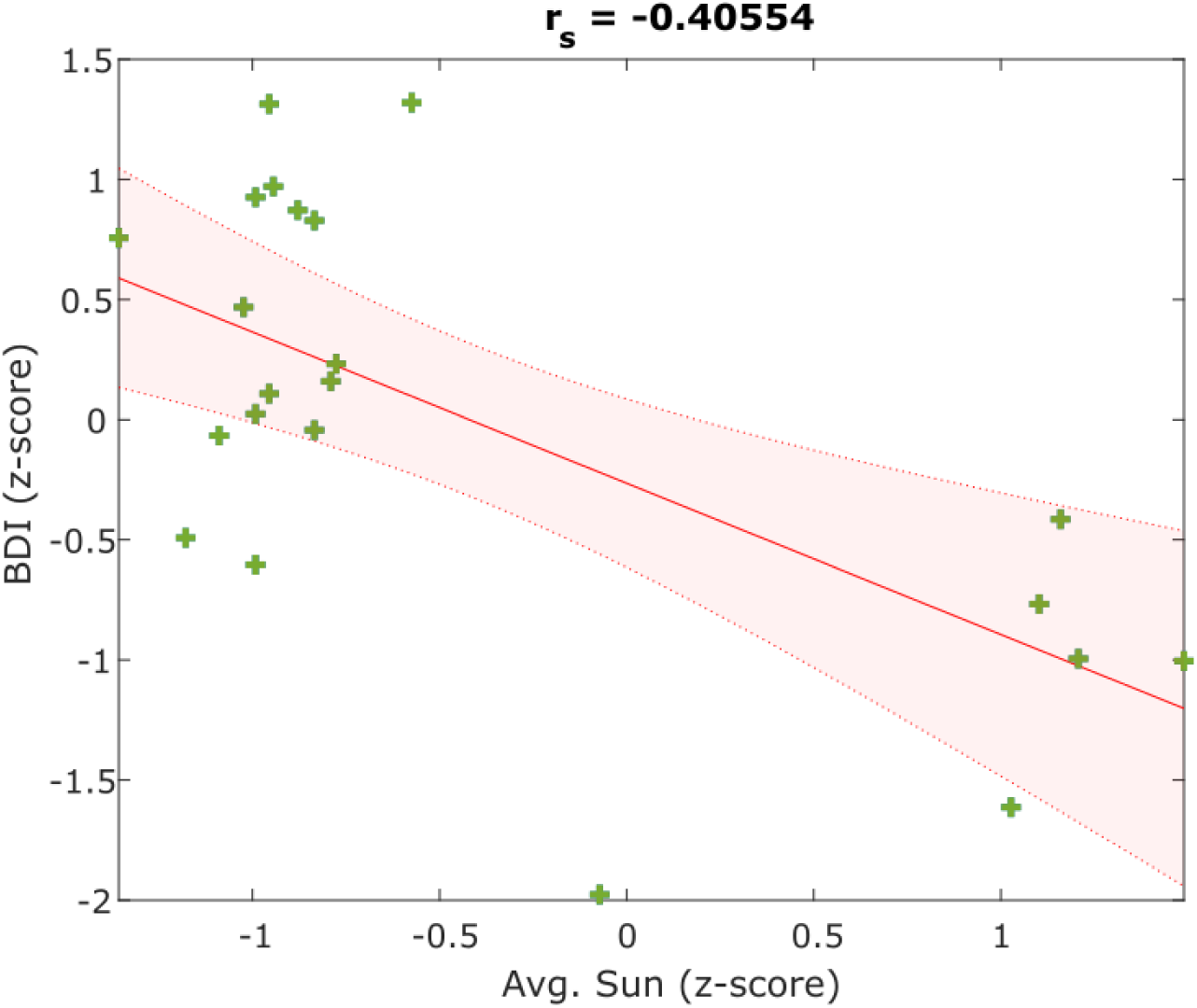
The plot shows the significant effect of average sunlight hours per day, averaged from a time frame of 28 days prior to blood sampling, on BDI scores in patients with SAD (p_uncorr_ < 0.01). Values are z-scored. An increase in average sunlight hours corresponds to a reduction in BDI scores, as confirmed by post hoc Spearman’s correlation analysis (r = −0.41).

## DISCUSSION

The study at hand explored the effect of average sunlight hours per day on *SLC6A4* promoter methylation in a cohort consisting of patients with SAD and HC. The analysis revealed that sunlight exposure significantly influenced *SLC6A4* promoter methylation, linking increased sunlight hours to reduced methylation levels of the promoter. The remaining covariates included in the main model (i.e., health status, 5-HTTLPR/rs25531 genotype, sex and age) did not show a significant effect on methylation levels. Our exploratory models examining the effect of average sunlight hours and *SLC6A4* promoter methylation on BDI scores indicated that average sunlight hours significantly influenced the BDI scores of patients with SAD, with more sunlight hours resulting in lower BDI scores. Furthermore, women (compared to men) exhibited elevated BDI scores, however this finding did not survive correction for multiple comparisons. *SLC6A4* promoter methylation did not exert a significant impact on BDI scores. The other factors assessed did not significantly affect BDI scores either.

### Analyses of *SLC6A4* Promoter Methylation

Epigenetic modifications influence how the DNA sequence is read to orchestrate protein synthesis. One form of epigenetic regulation is DNA methylation, with methyl groups attaching to specific sites on the DNA strand to regulate accessibility for RNA polymerases and thus gene activity. At specific CpG sites, elevated methylation leads to reduced transcriptional activity and thus lower protein levels (Lee, Teyssier et al. 2005). The gene *SLC6A4* codes for the SERT, whose relevance for SAD has been shown in numerous studies (Willeit, Sitte et al. 2008, Ruhe, Booij et al. 2009, Mc Mahon, Andersen et al. 2016, Tyrer, Levitan et al. 2016, Mc Mahon, Nørgaard et al. 2018). These investigations consistently suggest a diminished downregulation of SERT activity (i.e., elevated SERT activity) in patients with SAD during the winter months, resulting in increased serotonin reuptake and consequently lower serotonin availability. Nevertheless, epigenetic mechanisms potentially influencing SERT activity following a seasonal pattern have not yet been sufficiently investigated. DNA methylation per se is known to be influenced by meteorologic and seasonal factors (Bind, Zanobetti et al. 2014, Ricceri, Trevisan et al. 2014, Handschuh, Murgaš et al. 2023), including sunlight exposure (Aslibekyan, Dashti et al. 2014, Nair-Shalliker, Dhillon et al. 2014). Given the shown lack of downregulation of SERT density in patients with SAD during the winter months and the established impact of methylation on protein expression, it remains plausible to hypothesize that *SLC6A4* promoter methylation is influenced by seasonal variables, such as average daily sunlight duration. This would suggest a pattern of decreased methylation in winter, when sunlight hours are reduced, potentially leading to increased SERT expression and decreased serotonin availability, which may contribute to the development of winter-type depression. However, our findings indicated a different trend, suggesting that increased sunlight hours were associated with lower *SLC6A4* promoter methylation. This suggests that the relationship may be more nuanced, potentially influenced by additional factors beyond sunlight exposure alone, warranting further investigation. This trend is in fact in line with previously published findings, pointing towards reduced DNA methylation of certain genes after sun exposure when analyzing methylation in circulating peripheral blood cells, such as lymphocytes (Nair-Shalliker, Dhillon et al. 2014).

As described above, SERT is a crucial component in SAD pathophysiology, with higher SERT density during winter, resulting in worse clinical outcomes. Theoretically, lower *SLC6A4* promoter methylation levels could lead to increased transcriptional activity and protein synthesis, resulting in higher SERT density. Given that peripheral *SLC6A4* promoter methylation was higher (rather than lower) during periods of fewer sunlight hours per day (i.e., during autumn and winter), three theoretical conclusions can be drawn: First, the relationship between peripheral DNA methylation levels and brain SERT density, as well as neurotransmitter levels in SAD, may be more complex than initially assumed, suggesting that peripheral *SLC6A4* promoter methylation levels may not directly predict these variables in a straightforward or reliable manner. Second, it can be hypothesized that lower peripheral *SLC6A4* promoter methylation in response to more sunlight hours per day should be seen as a general effect of sunlight exposure found in various genes, as evidenced by previous methylation studies investigating the impact of sunlight on peripheral gene methylation (Aslibekyan, Dashti et al. 2014, Nair-Shalliker, Dhillon et al. 2014). Third, other biological mechanisms beyond sunlight exposure may be influencing *SLC6A4* promoter methylation, potentially overshadowing the direct effects of sunlight alone, suggesting that the relationship between sunlight hours and methylation may be part of a more complex network of regulatory processes, requiring further investigation to disentangle the relative contributions of these variables.

When examining the influence of peripheral DNA methylation on transcriptional processes related to *SLC6A4*, Wankerl et al. found that variations in SERT mRNA levels are unlikely to be influenced by DNA methylation patterns within the SERT gene (Wankerl, Miller et al. 2014). Another study published by Okada et al. reported that the methylation state of an *SLC6A4* CpG island (a gene region with elevated density of CpG dinucleotides) did not distinguish unmedicated patients diagnosed with major depression from either medicated patients or healthy participants (Okada, Morinobu et al. 2014), further questioning the relevance of average peripheral *SLC6A4* DNA methylation on transcription regulation. Conversely, Ouellet-Morin et al. found that increased *SLC6A4* DNA methylation was associated with bulling victimization when comparing the methylation state of bullied twins to the one of their non-bullied co-twins, supporting the hypothesis that childhood stress influences *SLC6A4* DNA methylation, thus linking methylation patterns to mental health conditions (Ouellet-Morin, Wong et al. 2013).

Mixed results in terms of risk factors for affective disorders modifying *SLC6A4* DNA methylation – with studies linking these risk factors to either higher (Zhao, Goldberg et al. 2013, Booij, Szyf et al. 2015, Duman and Canli 2015) or lower (Devlin, Brain et al. 2010, Alasaari, Lagus et al. 2012) methylation levels – also emphasize the complex interplay of *SLC6A4* DNA methylation, SERT expression, serotonin availability and environmental factors. As systematically reviewed by Provenzi et al., *SLC6A4* DNA methylation has been investigated in relation to various prenatal and postnatal adverse exposures, including maternal depression during pregnancy, perinatal and environmental stress as well as childhood trauma (Provenzi, Giorda et al. 2016). The authors found that *SLC6A4* might be seen as a relevant biomarker of early adversity exposures, with epigenetic mechanisms at this gene playing a critical role in programming.

Despite these efforts, establishing a causal relationship between *SLC6A4* DNA methylation and depression is challenging due to several factors inherent in the design and execution of human studies. First, the studies conducted so far employ different study designs, making direct comparisons difficult. Also, there is great variability in the specific methylation sites being analyzed, and further epigenetic mechanisms such as hydroxymethylation or histone modifications – potentially also influencing SERT expression – were not considered so far (Ell, Schiele et al. 2024). Additionally, study populations are rarely comparable regarding disease characteristics, medication use, age, and geographical location, further complicating the ability to draw definitive conclusions. Moreover, differences in sample size, statistical power, and the presence of confounding variables such as genetic background as well as pre- and postnatal stress and lifestyle factors contribute to the complexity (Schraut, Jakob et al. 2014). The dynamic nature of DNA methylation, influenced by both genetic and environmental factors over time, adds another layer of difficulty in establishing a direct causal link between *SLC6A4* DNA methylation and depression. Furthermore, as discussed earlier, we cannot be certain that *SLC6A4* promoter methylation data collected from peripheral blood samples accurately represents SERT density in the brain. In vivo imaging studies with clinical and healthy cohorts using, e.g., positron emission tomography, are needed to fill this gap in knowledge and provide a more accurate understanding of the relationship between peripheral *SLC6A4* DNA methylation, SERT density and clinical outcomes. Nevertheless, the study at hand is the first to analyze *SLC6A4* promoter methylation in SAD while considering meteorological data and the influence of sunlight on methylation dynamics. However, one limitation should be noted at this point: while we retrieved daily meteorological data for the 28 days prior to blood sampling, we did not monitor the exact locations of participants during this period, only that their primary residence was in Vienna. Despite this limitation, this approach is crucial for future investigations, as it underscores the potential impact of environmental factors on epigenetic modifications, offering a more comprehensive understanding of SAD. Such studies are relevant for clinical investigations in psychiatry because they can inform more targeted and effective treatments, considering both epigenetic and environmental influences on mental health.

### Beck Depression Inventory

BDI scores were used to assess the severity of depressive symptoms, allowing for the exploration of the potential relationship between clinical characteristics, sunlight exposure, and *SLC6A4* promoter methylation. Our exploratory models examining the effect of average sunlight hours and *SLC6A4* promoter methylation on BDI scores revealed several key findings. Average sunshine hours significantly influenced BDI scores in patients with SAD, with increased sunshine hours leading to lower BDI scores. This is consistent with previous clinical observations that sunlight exposure can alleviate depressive symptoms in SAD, possibly through its role in serotonin (Lambert, Reid et al. 2002) and vitamin D (Milaneschi, Hoogendijk et al. 2014, Akpınar and Karadağ 2022) synthesis, as well as in regulating circadian rhythms (Boivin, Duffy et al. 1996) by exerting positive effects on the suprachiasmatic nuclei (Bedrosian and Nelson 2017). Women had higher BDI scores than men in our study, although this finding was not significant after correction for multiple comparisons. The higher prevalence of depression in women (Hyde and Mezulis 2020) could be traced back to several factors, including hormonal fluctuations (Kundakovic and Rocks 2022), differential psychosocial stress management (Dong, Ironside et al. 2022) and gender-specific coping mechanisms (Graves, Hall et al. 2021). *SLC6A4* promoter methylation had no significant effect on BDI scores, nor did any of the other factors assessed. This suggests that while epigenetic modifications of *SLC6A4* may play a role in stress responses (Bakusic, Vrieze et al. 2020), their direct influence on depressive symptoms in SAD requires further investigation. Future studies should explore the interaction between (epi)genetic predisposition, environmental factors, and gender differences to better understand the complex mechanisms underlying SAD.

## Conclusion

The study at hand aimed to investigate the potential impact of average sunlight hours on *SLC6A4* promoter methylation and depression symptomatology in patients with SAD and healthy controls. Our findings demonstrated that increased sunlight exposure was significantly associated with reduced *SLC6A4* promoter methylation, suggesting an epigenetic response to seasonal changes of sunlight hours. Contrary to presumed expectations, *SLC6A4* promoter methylation increased with fewer sunlight hours per day, questioning the importance of *SLC6A4* DNA methylation in SAD. The observation that methylation was influenced solely by average sunlight hours, without a significant interaction with mental health status, suggests that this effect is rather general than pathophysiologically significant for SAD. Average sunlight hours were found to significantly influence BDI scores in patients with SAD, with more sunlight leading to lower BDI scores. This supports existing clinical observations that sunlight exposure can mitigate depressive symptoms. The lack of a significant impact of *SLC6A4* promoter methylation on BDI scores and other assessed factors highlights the complexity of the interplay between genetic, epigenetic, and environmental influences on SAD. These results indicate that the connection between sunlight exposure and methylation may be influenced by a more intricate set of regulatory mechanisms. The relationship between methylation and protein expression may be unclear or non-linear, and further research is needed to clarify the individual roles of these factors and to better understand how they interact in shaping methylation patterns.

Further research using in vivo imaging techniques, such as positron emission tomography (PET), is necessary to better understand the relationship between peripheral DNA methylation, brain SERT density, and clinical outcomes. This study is pioneering in its examination of *SLC6A4* promoter methylation in SAD while considering meteorological data, particularly sunlight exposure. This approach underscores the importance of environmental factors in epigenetic modifications and their potential role in psychiatric conditions.

## Data Availability

All data produced in the present study are available upon reasonable request to the authors.

## Contributors

M. Spies and R. Lanzenberger provided senior guidance, designed the main study and prepared the initial study protocol. D. Winkler and E. Winkler-Pjrek contributed valuable clinical and translational knowledge on SAD for the conceptualization of the main study. M. Spies and P. Baldinger-Melich were involved in clinical data collection. P. A. Handschuh and M. Spies designed the sub-study within the main study which was described in the section “Experimental Procedures”. A. M. Hartmann managed the analysis of genetic data. K. Domschke managed the analysis of epigenetic data. P. A. Handschuh performed the data collection from ©GeoSphere Austria (https://data.hub.geosphere.at), managed the literature research, and wrote the first draft of the manuscript. M. Murgaš undertook the statistical analysis and designed the figures. D. Rujescu provided valuable input on the coordination of the genetic analyses and facilitated communication between the research team and the laboratory. All authors contributed to and have approved the final manuscript.

## Acknowledgements

We thank M. Hienert, G. Gryglewski, T. Vanicek, A. Komorowski, A. Kautzky, L. Silberbauer J. Unterholzner, M.G. Godbersen, C. Kraus, and S. Kasper for clinical assistance; Ch. Ziegler for technical assistance in epigenetic analyses; G. Kranz, B. Spurny, S. Avramidis, and J. Jungwirth, J. Peters and K. Einenkel for administrative help.

